# Predicting risk of Alzheimer’s disease using polygenic risk scores developed for Parkinson’s disease

**DOI:** 10.1101/2023.08.16.23294135

**Authors:** Britney E. Graham, Scott M. Williams, Jason H. Moore

## Abstract

**Background and Objectives:** The two most common neurodegenerative diseases are Alzheimer’s disease (AD) and Parkinson’s disease (PD), both related to age and affect millions of people across the world, especially as life expectancy increases in certain countries. Here, we explore the potential predictiveness of the genetic risk of AD and PD separately and then the extent of the underlying shared genetics of AD and PD.

**Methods:** The population genetic risk estimates for AD and PD were derived using a previously developed population specific polygenic risk score (psPRS) and regression-based SNP filtering method. To test the overlap between AD and PD, we ran a regression of the AD psPRSs versus the population PD prevalences for both the filtered and unfiltered AD PRS and vice versa. We then assessed gene-gene interaction and pathway involvement using the Alzheimer’s KnowledgeBase (<u>AlzKB</u>) and STRINGdb, respectively.

**Results:** The unfiltered PD psPRS was moderately predictive, while the AD psPRS was not. After filtering, both the AD and PD, psPRSs improved to strongly predictive, explained most of the genetic variation. The ability of the unfiltered AD psPRS to predict PD, and vice versa, is poor. However, the filtered AD, and PD, psPRS were highly predictive.

**Discussion:** Our results suggest that there is a correlation between AD, and PD, specific allele frequency and prevalence, as well as an overlap of AD and PD generally. However, the AD psPRS is a better predictor of PD, than the PD psPRS is of AD. Our results call for further research into the general overlap of Alzheimer’s disease and Parkinson’s disease, despite the previous lack of evidence.

## Introduction

Neurodegenerative diseases affect millions of people across the world, and the two most common are Alzheimer’s disease (AD) and Parkinson’s disease (PD), both of which are age-related. Other neurodegenerative diseases include Huntington’s disease, Amyotrophic lateral sclerosis (ALS) and Motor neuron disease. All of these diseases are hallmarked by loss of nerve cell function in the brain or the peripheral nervous system^1^ and have some genetic overlap with each other^2^. These disorders, among other neurodegenerative diseases, are genetically complex with multiple causes, including genes and environment. Prevalence of these diseases varies among countries, due, possibly, to differences in socioeconomic status and life expectancy across populations. These diseases are becoming more prevalent everywhere as populations age. However, even age adjusted prevalence has increased, in PD more than in AD^3, 4^.

Identifying relevant risk factors for neurodegenerative diseases is important for early detection and prevention^5^. These diseases are relatively complex and have multiple components, including environmental and genetic. As multiple loci have been found that associate with these neurodegenerative diseases, it might be of use to combine these loci into a single genetic predictor. This can be done quantitatively by building a polygenic risk score (PRS).

### PRS

Polygenic risk scores (PRS) were developed with the intent of predicting diseases or phenotypes and usually incorporate, without interaction, multiple known risk associated loci. This follows the idea that effects are additive as opposed to multiplicative, i.e. epistatic. Often, the PRS is weighted by average estimated effects of risk alleles and, occasionally, the population allele frequency^6, 7^. However, as effect sizes can vary among populations, this method may limit generalizability^8^ and accurate prediction of understudied populations^9^. Modeling phenotypes with a PRS is more difficult when there is complex genetic architecture and environmental factors^10^.

Disease risk prediction using genetic factors is usually done for individuals. Examples of PRS use include predicting obesity and type 2 diabetes mellites, various cancers and heart conditions, and IQ and mental illness at the individual level using population-derived associations. However, although population-level prevalence prediction is relatively new, we have previously shown that population-level, risk allele frequency-based, polygenic risk scores (PRSs) can explain prevalence variation, to varying degrees, among geographic populations^11^.

PRSs have been used in both AD and PD in the past but have mostly been able to account for only a small amount of heritability, although, there is some evidence that AD PRSs are transferable across populations^12^. Here, using our previous method, we explore the potential predictiveness of the genetic risk of AD and PD separately^11^. We then attempt to explain the extent of the underlying shared genetics of AD and PD.

### AD-PD Similarities

Although AD and PD are distinct diseases, they have many similarities. Neither AD, nor PD, are curable and only palliative treatments are available. Both are progressive, neurodegenerative, related to advanced age, characterized by protein buildup in the brain and affect movement and cognition. Olfactory impairment is an early symptom in both AD and PD^13^

Despite phenotypic similarities, AD and PD are rarely co-morbid^14^, but, interestingly, family history of AD increases the risk of developing PD and vice versa^15, 16^.While there has been indication of only minor genetic overlap between AD and PD, both AD and PD share differentially expressed genes and pathways^17^. Indeed, the KEGG PD pathway has a 78.2% overlap with the KEGG AD pathway^18^. They also share some disease mechanisms^19^, treatments^20^ and environmental risk factors^21^

### Alzheimer’s disease

First described in 1906 by Alois Alzheimer^22^, Alzheimer’s disease (AD), the most common neurodegenerative disease, is marked by increasing loss of memory, reasoning and bodily functions, leading to death. There are two main forms of AD. The most common is late onset (LOAD, 95% of cases), where patients are diagnosed in their mid-60s or older with an average survival time after diagnosis of four to eight years. LOAD is, notably, more common in women^23^.

Treatments for AD are palliative, except for aducanumab (targets the amyloid beta protein), which has had mixed results^24^. Aside from age, risk factors include vascular disease, family history of AD, mid-life hypertension, obesity and diabetes^23^. While AD does have an environmental component, AD heritability, estimated by twin studies, is 58%-79%^25^. However, even when the strongest genetic risk factor for LOAD, *APOE ε4* allele^26^, is combined with other known genetic factors, the variance explained is still only 24–33%^27^.

There is an unequal distribution of AD worldwide, even taking the average age of a population into account^3^. Western Europe has the highest prevalence of AD and dementia and Sub-Saharan Africa the lowest. The fastest increase in population AD prevalence is in China, India, and other south Asian and western Pacific countries^3^. This is especially impactful as the proportion of people over 65, and cost of care, increases across the world and the size of the workforce decreases in some areas^28^.

### Parkinson’s disease

Parkinson’s disease (PD), first described in 1817 by James Parkinson^29^, is the second most common neurodegenerative disorder after AD^1^. PD has a range of symptoms including uncontrollable movement, shaking, difficulty speaking and walking As with AD, PD is usually diagnosed after the age of 60^30^, with only 3–5% of cases occurring earlier^31^. Contrasting with AD, PD occurs more frequently in men than in women^32^. Although PD tends to primarily affect the motor system, 20% to 40% of PD patients develop PD specific dementia, but only after the onset of the other symptoms^33^.

There is no cure for PD, but there are a variety of palliative treatments for symptoms, including a range of drugs, primarily levodopa^34^. Occupational, physical, and speech therapy have been used to help with balance, gait speed and voice and speech disorders. Surgeries to achieve deep brain stimulation have been somewhat successful in improving motor function and help to decrease motor fluctuations, dyskinesia, and antiparkinsonian medication use^35^.

While the actual cause of PD is still uncertain, there seem to be both environmental and genetic risk factors^36^. The heritable portion of PD is estimated to be 22%^36^, but only about 16-36% of that heritability has been explained^37^. Confirmed dominant genes include *SNCA, LRRK2, GBA*, and *VPS35*, and recessive genes include *Parkin, PINK1, DJ1*. Prevalence of PD is higher in people of European decent and appears to be lower in Africa^4^. However, prevalence data by ethnicity is sometime contradictory^38^.

The goal of this study was to assess the overlap of PRS components for AD and PD and the cross-predictive value. We did this by assessing to what extent PRSs could predict population prevalence of each disease, and then each disease PRS for the other.

## Materials and Methods

### Allele and Prevalence Data Collection

We assessed the role of PRSs in predicting population prevalence among the populations included in the International Genome Sample Resource (IGSR) from the 1000 Genomes Project^39^. Prevalence data for each population in the 1000 Genomes (Table 1) was collected from the literature, principally from country level data^3^. Using a threshold of p < 1 × 10^−5^, we extracted AD associated alleles from the GWAS catalog using Alzheimer’s disease as a search term (MONDO_0004975). We then mined the allele frequencies for each SNP from the 1000 Genomes project for each population (Table S1).

**Table 1.** Relative Alzheimer’s and Parkinson’s disease prevalence among populations.

|  |  |  |  |
| --- | --- | --- | --- |
| 1000 Genomes Super population | 1000 Genomes Population | Alzheimer's disease prevalence<sup>3</sup> | Parkinson's disease prevalence<sup>4</sup> |
| African (AFR) | African Caribbean in Barbados (ACB) | $7.78 \times 10^{-3}$ | |
| | Americans of African Ancestry in SW USA (ASW) | $1.37 \times 10^{-2}$ | $1.04 \times 10^{-2}$ |
| | Esan in Nigeria (ESN) | $8.21 \times 10^{-4}$ | |
| | Gambian in Western Divisions in the Gambia (GWD) | $7.89 \times 10^{-4}$ | $8.65 \times 10^{-5}$ |
| | Luhya in Webuye, Kenya (LWK) | $1.25 \times 10^{-3}$ | $1.34 \times 10^{-4}$ |
| | Mende in Sierra Leone (MSL) | $7.66 \times 10^{-4}$ | $8.69 \times 10^{-5}$ |
| | Yoruba in Ibadan, Nigeria (YRI) | $8.21 \times 10^{-4}$ | $1.12 \times 10^{-4}$ |
| Ad Mixed American (AMR) | Colombians from Medellin, Colombia (CLM) | $5.16 \times 10^{-3}$ | $5.38 \times 10^{-4}$ |
| | Mexican Ancestry from Los Angeles USA (MXL) | $5.51 \times 10^{-3}$ | $5.57 \times 10^{-4}$ |
| | Peruvians from Lima, Peru (PEL) | $3.49 \times 10^{-3}$ | $5.47 \times 10^{-4}$ |
| | Puerto Ricans from Puerto Rico (PUR) | $9.50 \times 10^{-3}$ | $1.26 \times 10^{-3}$ |
| East Asian (EAS) | Chinese Dai in Xishuangbanna, China (CDX) | $7.37 \times 10^{-3}$ | $1.01 \times 10^{-3}$ |
| | Han Chinese in Beijing, China (CHB) | $7.37 \times 10^{-3}$ | $1.01 \times 10^{-3}$ |
| | Southern Han Chinese (CHS) | $7.37 \times 10^{-3}$ | $1.01 \times 10^{-3}$ |
| | Japanese in Tokyo, Japan (JPT) | $2.76 \times 10^{-2}$ | $2.02 \times 10^{-3}$ |
| | Kinh in Ho Chi Minh City, Vietnam (KHV) | $7.00 \times 10^{-3}$ | $6.88 \times 10^{-4}$ |
| European (EUR) | Utah Residents (CEPH) with Northern and Western European Ancestry (CEU) | $1.69 \times 10^{-2}$ | $1.67 \times 10^{-2}$ |
| | Finnish in Finland (FIN) | $1.53 \times 10^{-2}$ | $1.87 \times 10^{-3}$ |
| | British in England and Scotland (GBR) | $1.27 \times 10^{-2}$ | $1.77 \times 10^{-3}$ |
| | Iberian Population in Spain (IBS) | $1.78 \times 10^{-2}$ | $2.00 \times 10^{-3}$ |
| | Toscani in Italia (TSI) | $2.26 \times 10^{-2}$ | $2.39 \times 10^{-3}$ |
| South Asian (SAS) | Bengali from Bangladesh (BEB) | $2.54 \times 10^{-3}$ | $3.43 \times 10^{-4}$ |
| | Gujarati Indian from Houston, Texas (GIH) | $2.22 \times 10^{-3}$ | $4.35 \times 10^{-4}$ |
| | Indian Telugu from the UK (ITU) | $2.22 \times 10^{-3}$ | $4.35 \times 10^{-4}$ |
| | Punjabi from Lahore, Pakistan (PJL) | $1.70 \times 10^{-3}$ | $2.76 \times 10^{-4}$ |
| | Sri Lankan Tamil from the UK (STU) | $6.06 \times 10^{-3}$ | $8.40 \times 10^{-4}$ |

For the prediction of PD, we followed the same procedure as for AD. Using Parkinson’s disease as a search term (MONDO_0005180), we extracted the associated alleles at a threshold of p < 1 × 10^−5^ from the GWAS catalog. The corresponding allele frequencies were taken from the 1000 Genomes project for each population. Additionally, we collected prevalence data for PD from literature^4^.

### Polygenic risk scores

The population genetic risk estimate was derived using a previously developed population specific polygenic risk score (psPRS)^11^. This method uses the frequency of all disease risk alleles for a population to predict that population’s disease prevalence. As effect size is sometimes unavailable and usually only based on a limited number of populations, the psPRS uses only the population risk allele frequencies. Using the simplest assumption that the genetic architecture of the phenotype of interest is additive, the psPRS was a summation of the population risk allele frequencies in each population. For a population in the 1000 Genomes database, psPRS is the PRS for that population and *p*_*i*_ is the allele frequency of *SNP*_*i*_:

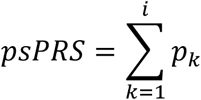

We then performed regression analyses between the psPRSs and population prevalences, establishing the relationship between the psPRS and the population prevalence of each disease.

#### Maximization of the coefficient of determination sensitivity analysis

To obtain an optimal and likely universal PRS, we then filtered the risk associated SNPs based on a maximization of the coefficient of determination (r^2^) of the regression of the psPRS with the population prevalence. We eliminated SNPs that reduced the predictiveness of each psPRS by assessing the effect of removing individual SNPs from the psPRS.

As an example, the equation for the sensitivity analysis for *SNP*_*2*_ would be:

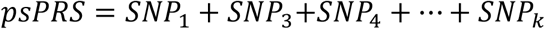

Each SNP was annotated with the r^2^ value calculated for the linear regression between the psPRS, without that SNP, and the population disease prevalence. SNPs that negatively affected the predictability of the model were removed. We then repeated this process until only the most predictive SNPs remained. Assuming additivity, we expect the model with the largest r^2^ to include all true disease associating SNPs with universal effects, i.e., those that align with disease prevalence across a broad array of populations. This method, as well as the psPRS algorithm, was described in our previous work^11^.

#### Intersection

Given the known similarities between AD and PD, we sought to determine if AD and PD are similar enough genetically that the AD PRS would be able to predict PD population prevalence and vice versa. We ran a regression of the AD psPRSs versus the population PD prevalences for both the filtered and unfiltered AD PRS. We then ran the PD psPRS versus the population AD prevalences for both the filtered and unfiltered PD PRS.

#### Gene-Gene interaction

To identify possible genetic links between AD and PD, we explored the underlying interaction of the AD and PD specific genes using the Alzheimer’s KnowledgeBase (<u>AlzKB</u>), a graph database that incorporates various AD resources into one graph. The AlzKB nodes include genes, pathways, drugs, and diseases, and the edges represent various relationships between nodes, including, but not limited to, co-expression, chemical effect and gene-gene interaction. We queried the AlzKB with the genes that have been associated with PD in several studies^40-42^, and determined which of the AD genes interacted directly with the PD genes, based on the relationship term in the graph (GENEINTERACTSWITHGENE), which is in turn based on protein-protein interaction (PPI)^43^.

#### Pathway analysis

We then assessed the possible pathway involvement of the interacting AD and PD genes from AlzKB using the STRING database of Protein-Protein Interaction Networks Functional Enrichment Analysis. STRINGdb does not just contain PPI networks, but also incorporates enrichment analysis using other databases, including Gene Ontology and KEGG^44^.

## Results

### Alzheimer’s Disease

There are 1386 AD-associated SNPs in the GWAS catalog (Table S1). The AD psPRS using these SNPs had a positive but non-significant relationship, with AD prevalence (Figure 1A, slope: 0.0005, R²: 0.11, p-value: 0.091). Intracontinental populations similarly had weak, non-significant relationships, apart from the south Asian and Ad Mixed American population clusters (Figure S1A). Both superpopulations had a moderate r^2^ value; 0.49 and 0.42, respectively, although still without a significant p-value. However, the south Asian population cluster showed a negative relationship between the psPRS and AD prevalence.

**Figure 1.**
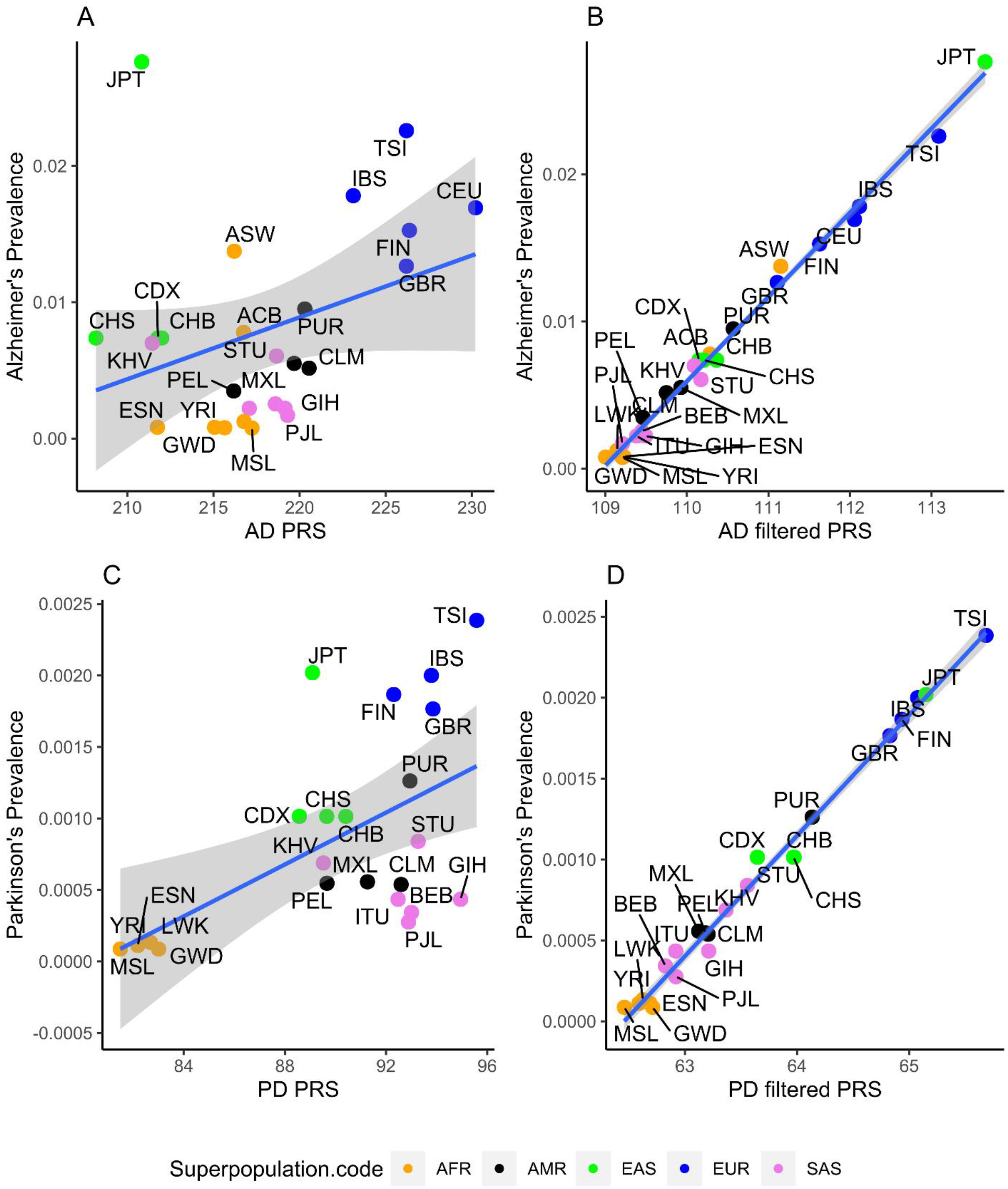
Correlation between diseases and the corresponding psPRSs. The data points are colored according to the super populations: AFR (orange), AMR (black), EAS (green), EUR (blue) and SAS (purple). The scale of the x-axis is not the same for both plots due to differing psPRS ranges. **A**. Full AD vs AD PRS model (r^2^ =0.11; *p*-value: 0.091). **B**. After maximization (r^2^ = 0.99, *p*-value: < 2.2 × 10^−16^). **C**. Correlation between Parkinson’s Disease and the PD full model psPRS (r^2^ =0.33; *p*-value: 0.0045). **D**. After maximization (r^2^ = 0.99, *p*-value: < 2.2 × 10^−16^).

After maximization filtering, 387 SNPs remained in the PRS model (Table S2). The relationship between the filtered psPRSs and AD was strong, positive and highly significant (Figure 1B, Slope: 0.0057, r²: 0.99, p-value: < 2.2 × 10^−16^) (Figure 1B). Intracontinental populations also showed the same patterns of significance and strength, although the south Asian-AD relationship switched direction from negative to positive (Figure S1B).

### Parkinson’s disease

There were 450 PD associated SNPs in the GWAS catalog (Table S3). The PD psPRS using these SNPs had a significant association with PD prevalence (Figure 1C.,slope: 9 × 10^−5^, r²: 0.33, p-value: 0.0045). The correlation between the PD psPRSs and the PD prevalences was best in the European superpopulation (r² = 0.64) but was not statistically significant (Figure S2A; p-value: 0.20).

After maximization filtering, 242 SNPs remained (Table S4), yielding a positive, very significant and predictive, relationship (Figure 1D, slope = 0.0007, r² = 0.99, p-value: < 2.2 × 10^−16^). The superpopulation correlations improved in general, with the European populations, again, performing the best (Figure S2B, r² = 0.99, p-value: 0.0044).

### AD and PD Intersection

The regression analysis between the unfiltered AD psPRSs and PD prevalences showed a slightly positive, but only somewhat, predictive model (Figure 2A, slope: 6 × 10^−5^, r²: 0.17, p-value: 0.052). Intracontinental populations mostly had weak, albeit non-significant relationships, apart from the European population cluster, which showed a strong, though non-significant r^2^ value of 0.71. Additionally, the south Asian population cluster again showed a negative relationship between the psPRS and PD prevalence (Figure S3A).

**Figure 2.**
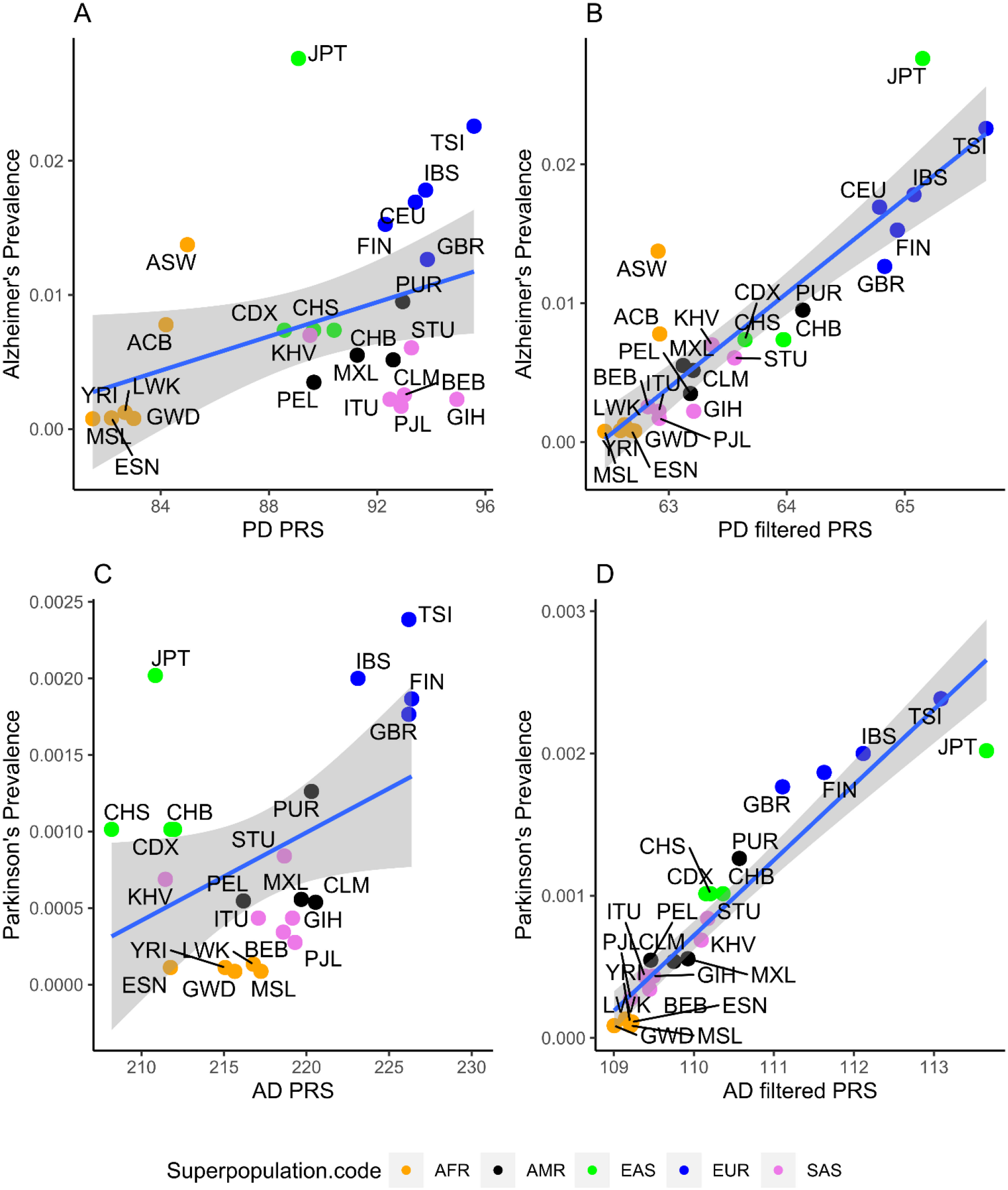
Correlation between diseases and the exchanged psPRS. The data points are colored according to the super populations: AFR (orange), AMR (black), EAS (green), EUR (blue) and SAS (purple). The scale of the x-axis is not the same for both plots due to differing psPRS ranges. **A**. Full PD prevalence and the AD psPRS model (r^2^ = 0.17; p-value: 0.052). **B**. After maximization (r^2^ = 0.90, p-value: 3.56 × 10-12). **C**. AD prevalence and the PD psPRS full model (r^2^ =0.16; *p*-value: 0.045). **D**. After maximization (r^2^ =0.79, *p*-value: 1.41 × 10^−9^).

When the same analysis was performed with the filtered AD psPRS values, the PD relationship was both highly significant and predictive (Figure 2B, slope: 0.0005, r² = 0.90, p-value: 3.56e-12). However, while most of the continental population clusters showed an improvement in strength and significance of the psPRS versus prevalence relationship, the European populations experienced a reduction of both significance and strength from the unfiltered comparison (Figure S3B).

The regression analysis between the unfiltered PD psPRSs and AD prevalences showed a slightly positive, significant, predictive model (Figure 2C and S4A, slope: 0.0006, r²: 0.16, p-value: 0.045). When the same analysis was performed with the filtered PD psPRS values, the AD relationship improved considerably in significance and predictiveness (Figure 2D and S4B, slope: 0.0068, r² = 0.79, p-value: 1.41 × 10^−9^).

### AD and PD gene-gene interaction and pathway analysis

Interestingly, only two PD genes, *MAPT* and *HLA-DRB5*, were directly connected to AD in AlzKB. However, there were several interactions in AlzKB between the PD gene set and AD gene set (Figure 3, Table 2), but only *MAPT* is in the intersection of the two sets and interacts with 11 genes in total. In addition, we used STRINGdb to identify enrichment of *CASP3, ATP5F1A, SNCA* and *MAPT* (FDR: 6.14 × 10^−7^) in the Kegg AD and PD pathways (Figure 4). *CASP3* and *ATP5F1A* are also in other neurodegenerative KEGG pathways, namely Huntington’s disease and Amyotrophic Lateral Sclerosis. In fact, it has been observed that 91% of KEGG pathways are potentially connected to AD pathogenesis^45^.

**Table 2.** Interacting Alzheimer’s and Parkinson’s genes in AlzKB.

| **Table 2.** Interacting Alzheimer's and Parkinson's genes in AlzKB. |  |
| --- | --- |
| Alzheimer's genes | Parkinson's genes |
| APOE | MAPT |
| APP | SNCA, GAK, RPS12, TRIM40, UCHL1, DDRGK1, MAPT, HTRA2, VPS35 |
| ATP5F1A | RPS12 |
| BAX | SNCA |
| BIN1 | SYNJ1, ATXN2 |
| CALM1 | PAM, MAPT, DNAJC13, SNCA |
| CASP3 | MAPT, FYN, DCTN1, KCNIP3 |
| CD2AP | FYN |
| CHRNA7 | FYN |
| ENO1 | FYN, RPS12 |
| ESR1 | EIF4G1, UBTF, RPS12, GAK, TBP |
| GSK3B | DCTN1, MAPT, SNCA, DNAJC13 |
| IGF1R | KCNIP3 |
| IL1B | TBP |
| MAPT | DCTN1, FYN, PRKN, LRRK2, SNCA |
| PLCG2 | CD19, FYN |
| PSEN1 | MAPT, HTRA2, SCAF11, KCNIP3 |
| PSEN2 | KCNIP3 |
| VEGFA | VPS35 |

**Figure 3.**
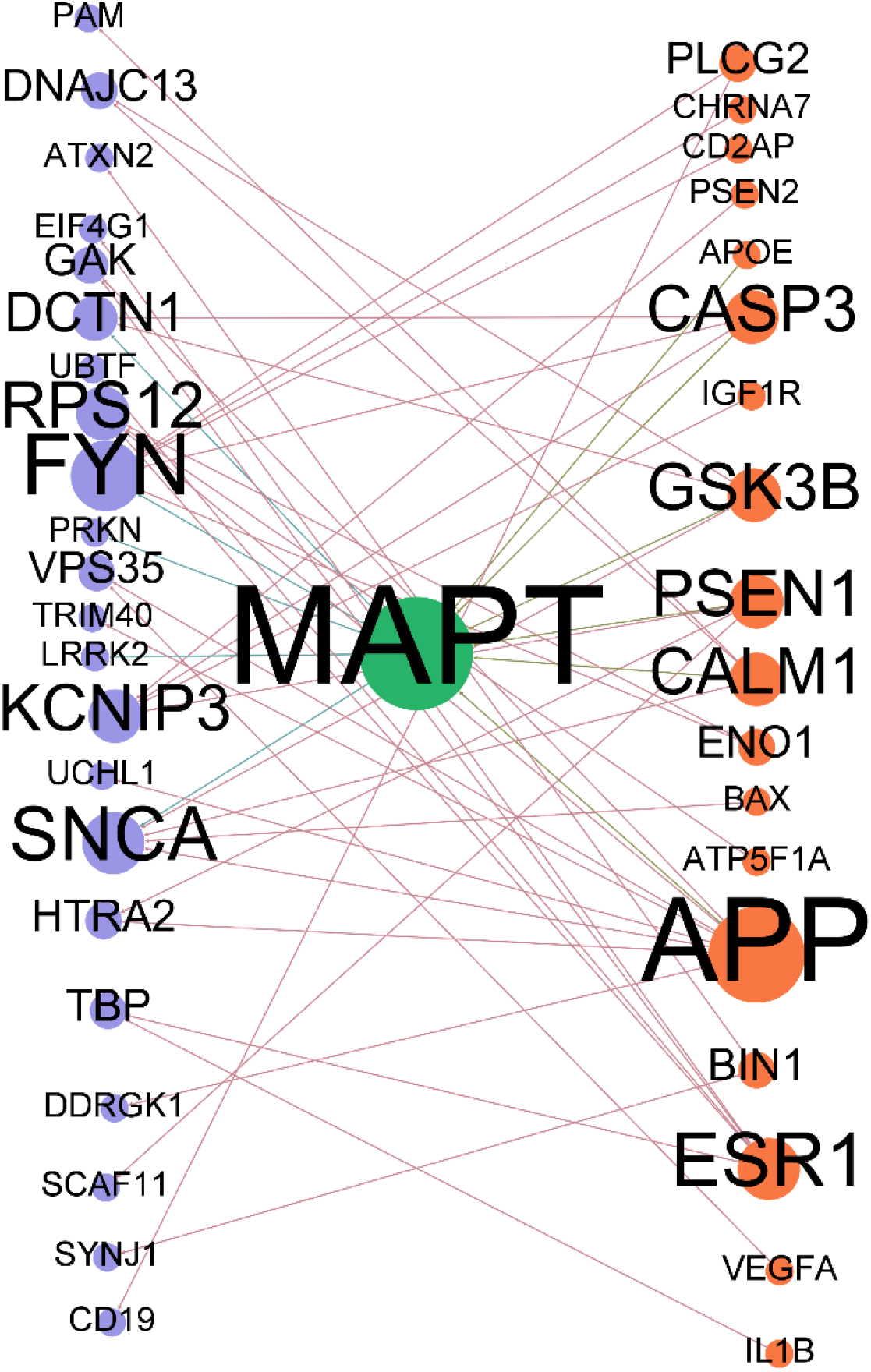
Bipartite network of AD gene and PD gene interactions from AlzKB. Interactions in AlzKB between the PD gene set (blue) and AD gene set (orange), but only *MAPT* (green) is in the intersection of the two sets and interacts with 11 genes in total.

**Figure 4.**
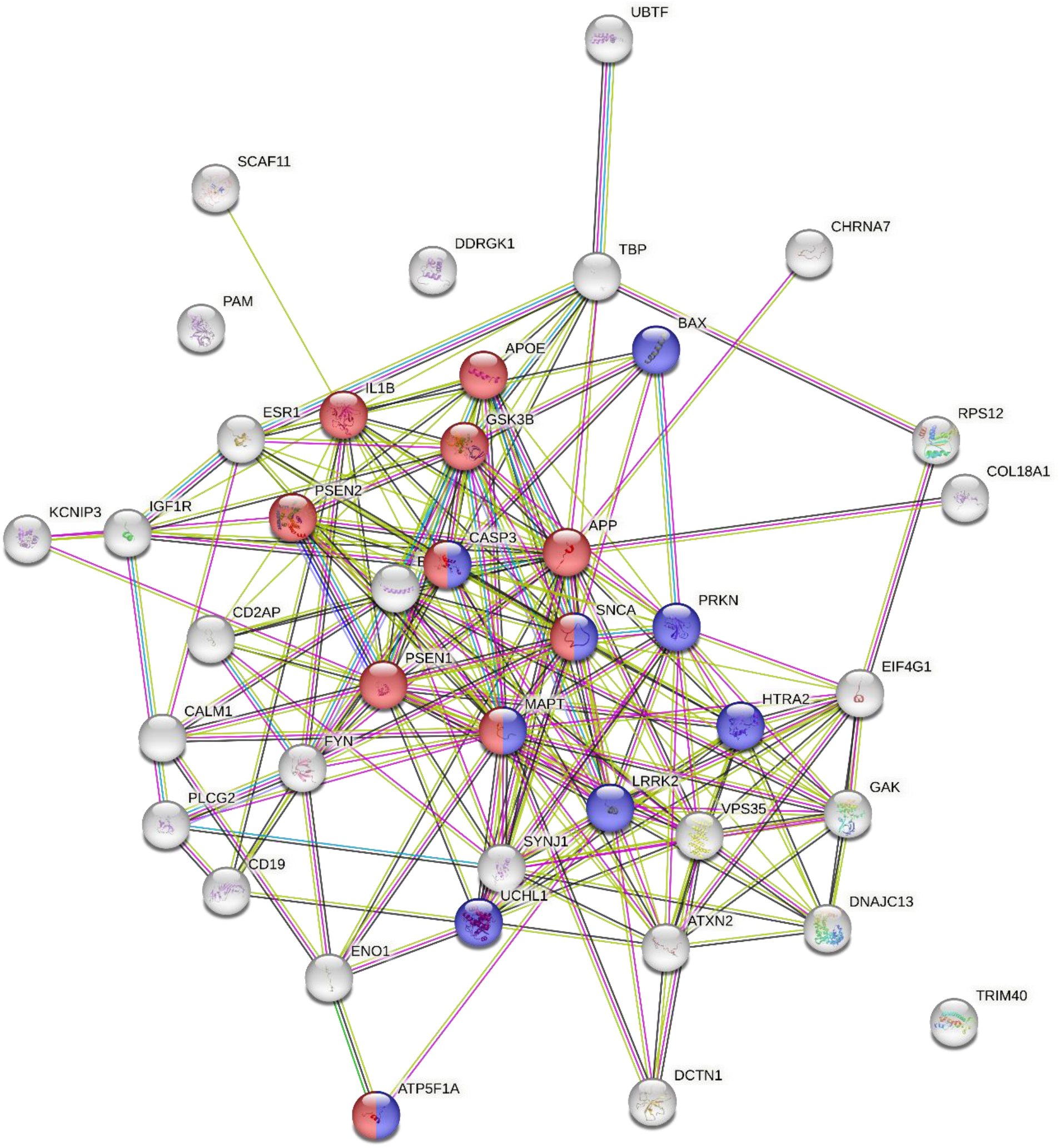
STRINGdb enrichment of *CASP3, ATP5F1A, SNCA* and *MAPT* in the Kegg AD (blue) and PD (red) pathways.

## Discussion/Conclusion

### AD

While the full unpruned relationship between the psPRS and AD prevalence was not significant, our pruned results were, suggesting that there is a correlation between AD specific allele frequency and prevalence. That the variance explained was low, supports an environmental and/or interactive piece of the overall etiology of AD. However, the significance of the filtered results suggests that there is a specific subset of AD loci that can predict the prevalence of AD with high accuracy in most populations.

### AD-PD analyses

That the unfiltered AD PRS can, with statistical significance, predict the prevalence of PD in certain populations indicate that there may be a substantial shared underlying genetic risk for both diseases. As the AD specific filtering also improved the PD-AD-PRS comparison, it seems that same subset of AD loci can predict the prevalence of PD with high accuracy in many populations. Interestingly, the European population comparisons exchanged favorability between the filtered and unfiltered PRS. The strength and significance of AD unfiltered PRS-prevalence relationship was far less than the PD. The relationship for PD worsened with the filtering, possibly because African and European American PD prevalences are much higher than their ancestral groups. Perhaps this is due to different environmental conditions in the United States, than in ancestral lands.

This genetic link between AD and PD is further underlined by our analysis of the relationship between the PD psPRS and AD prevalence. Between the AD PRS and the PD PRSs ability to predict AD prevalence, the best predictor over all is the filtered AD PRS. Yet, the PD PRS is at least significant and somewhat predictive before filtering and even more so after. The ability of the AD PRS to predict PD is still better than the ability of the PD PRS to predict AD. However, that the filtered PD PRS r^2^ is so high, still indicates a, as yet unidentified, strong underlying interdependency of PD and AD.

Although AD and PD are distinct diseases, these results suggest that they are genetically similar enough that a PRS developed for one disease can be applied to the other to estimate population disease risk. This is supported by the gene-gene interaction observed in our AlzKB based analysis of the AD and PD specific genes. As only two genes, *MAPT* and *HLA-DRB5* are shared between AD and PD directly, this suggests that either there are shared genes yet to be characterized or the underlying common genetics are significantly more complex than previously thought.

The four AlzKB gene overlap of the KEGG AD and PD pathways shows a connection beyond AD and PD simply sharing two genes directly. The additional overlap with the Amyotrophic Lateral Sclerosis and Huntington’s disease KEGG pathways, underlines the neurodegenerative role of these genes. Additionally, the 78.2% overlap of the full KEGG PD pathway with the full AD pathway supports a significant underlying genetic and functional connection between AD and PD in general.

Our analyses shows that *MAPT* is an important gene in both AD and PD and may be the key for understanding the overlap of AD and PD. Indeed, *MAPT* is involved in tau and is primarily expressed in the brain^46^. *MAPT* is then a candidate for gene therapy in downregulating tau protein^47^. That *MAPT* is so central may not be surprising, as it has been suggested as a supergene ^48, 49^.

One thing to note, is that AD and PD prevalences are somewhat correlated in the 1000 Genomes populations. Indeed, the prevalences of Dementia, Parkinson’s disease, multiple sclerosis, and motor neuron diseases are highly correlated worldwide^50^. A regression analysis of the PD prevalences versus the AD prevalences shows a statistically significant moderate positive correlation (Slope: 0.23, p-value: 0.018, r^2^: 0.47). Therefore, we cannot rule out that this perceived link between AD and PD is based on shared environmental differences between populations. In conclusion, we have shown evidence supporting significant shared underlying biology for AD and PD, although it has continued to be elusive in other studies. Our results call for further research into the general overlap of Alzheimer’s disease and Parkinson’s disease, despite the previous lack of evidence.

## Data Availability

All data produced in the present work are contained in the manuscript.

