## Supplemental Figures for "Predicting risk of Alzheimer’s disease using polygenic risk scores developed for Parkinson’s disease"

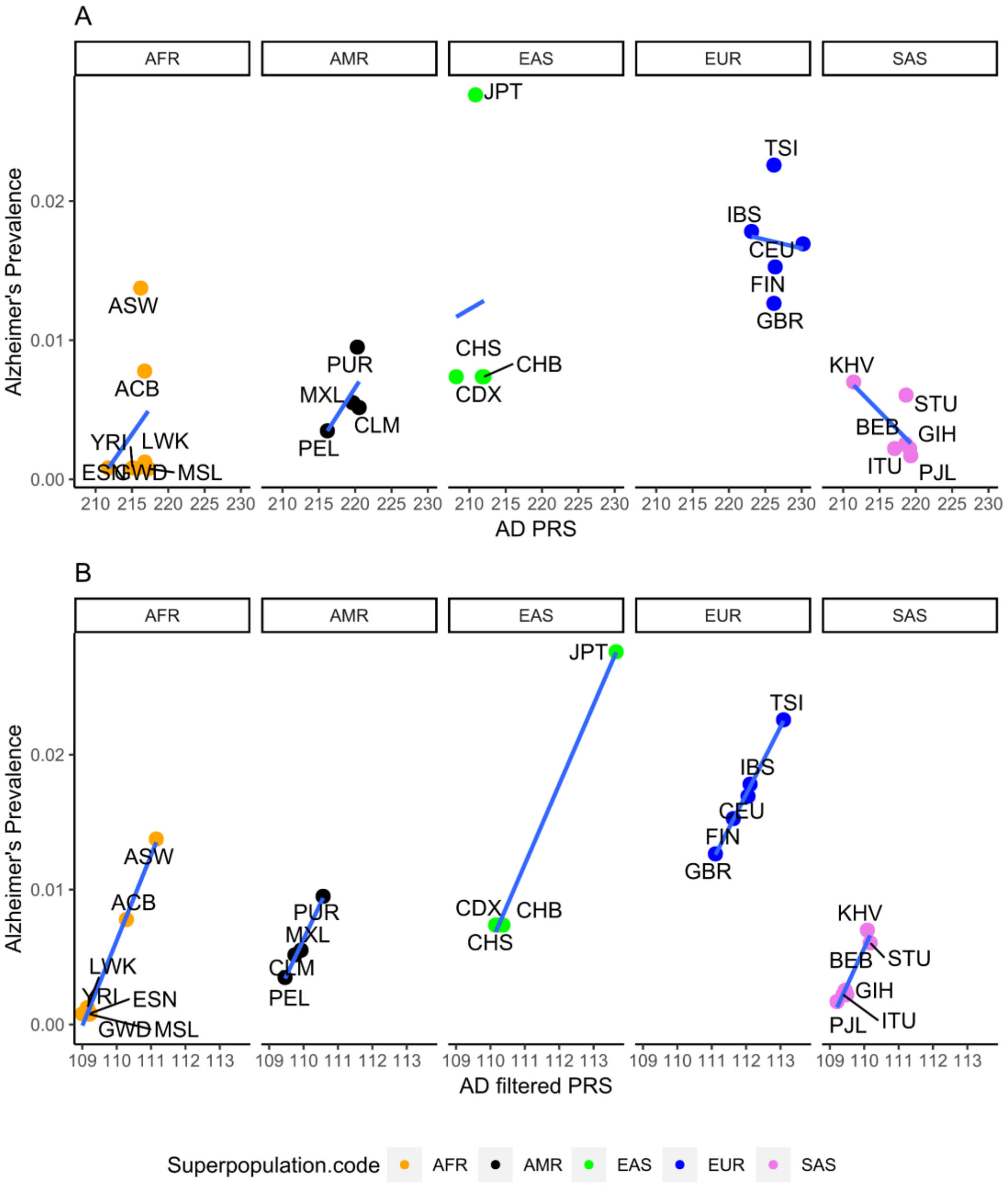


**Figure S1.** AD PRS versus AD prevalence separated by super population. The data points are colored according to the super populations: AFR (orange), AMR (black), EAS (green), EUR (blue) and SAS (purple). A) Full model by super population. B) Super populations after maximization.

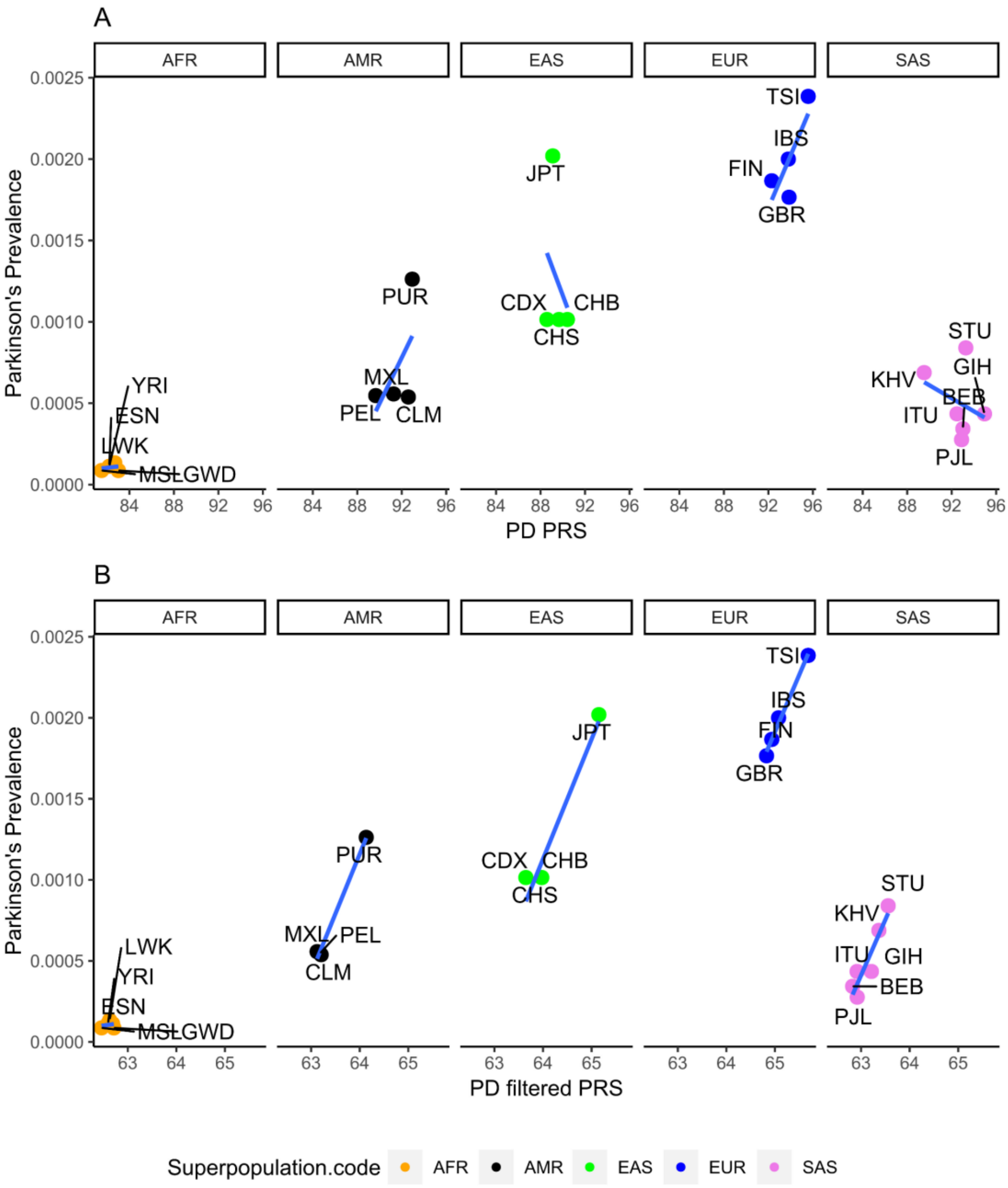


**Figure S2.** PD PRS versus PD prevalence separated by super population. The data points are colored according to the super populations: AFR (orange), AMR (black), EAS (green), EUR (blue) and SAS (purple). A) Full model by super population. B) Super populations after maximization.

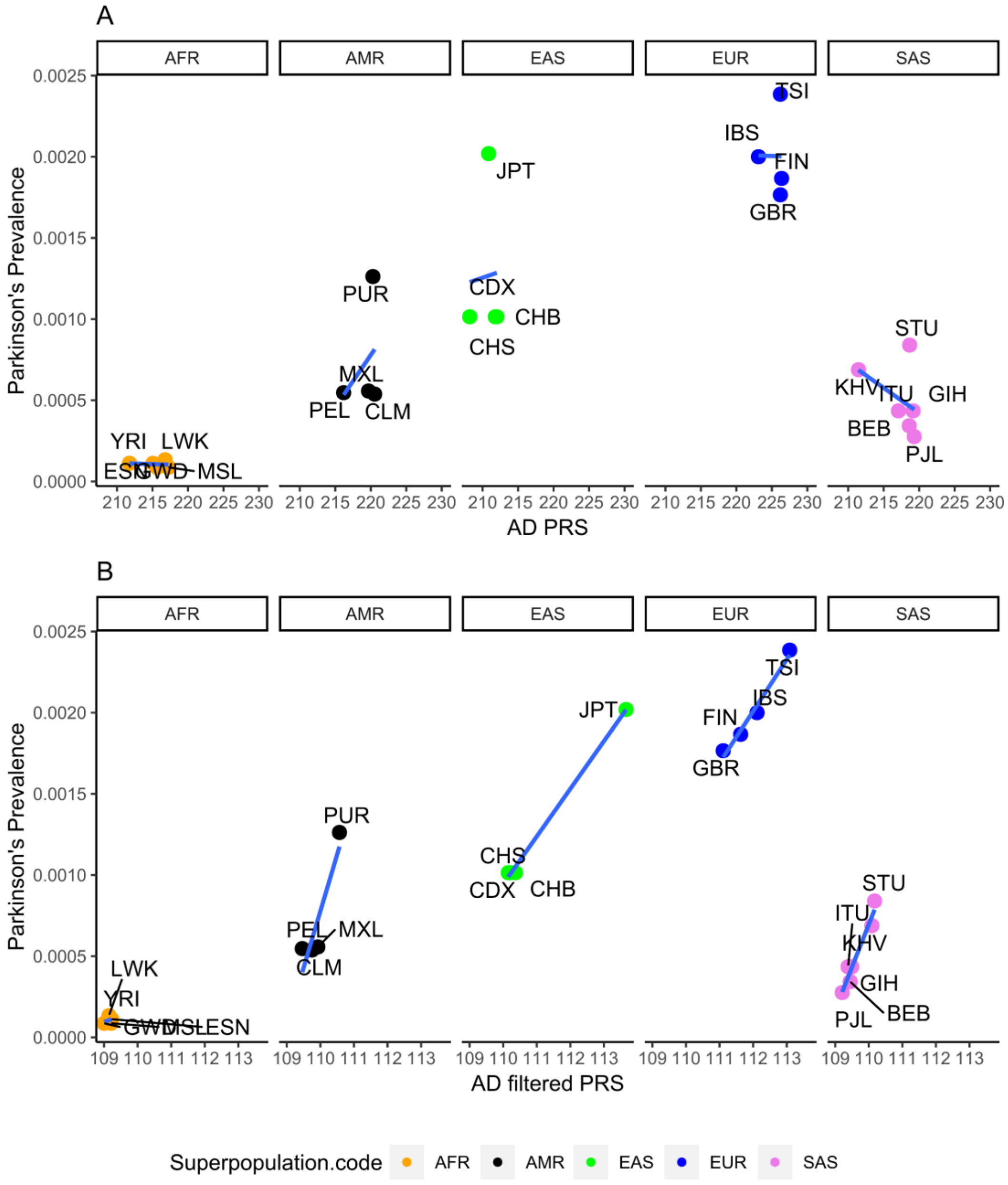


**Figure S3.** AD PRS versus PD prevalence separated by super population. The data points are colored according to the super populations: AFR (orange), AMR (black), EAS (green), EUR (blue) and SAS (purple). A) Full model by super population. B) Super populations after maximization.

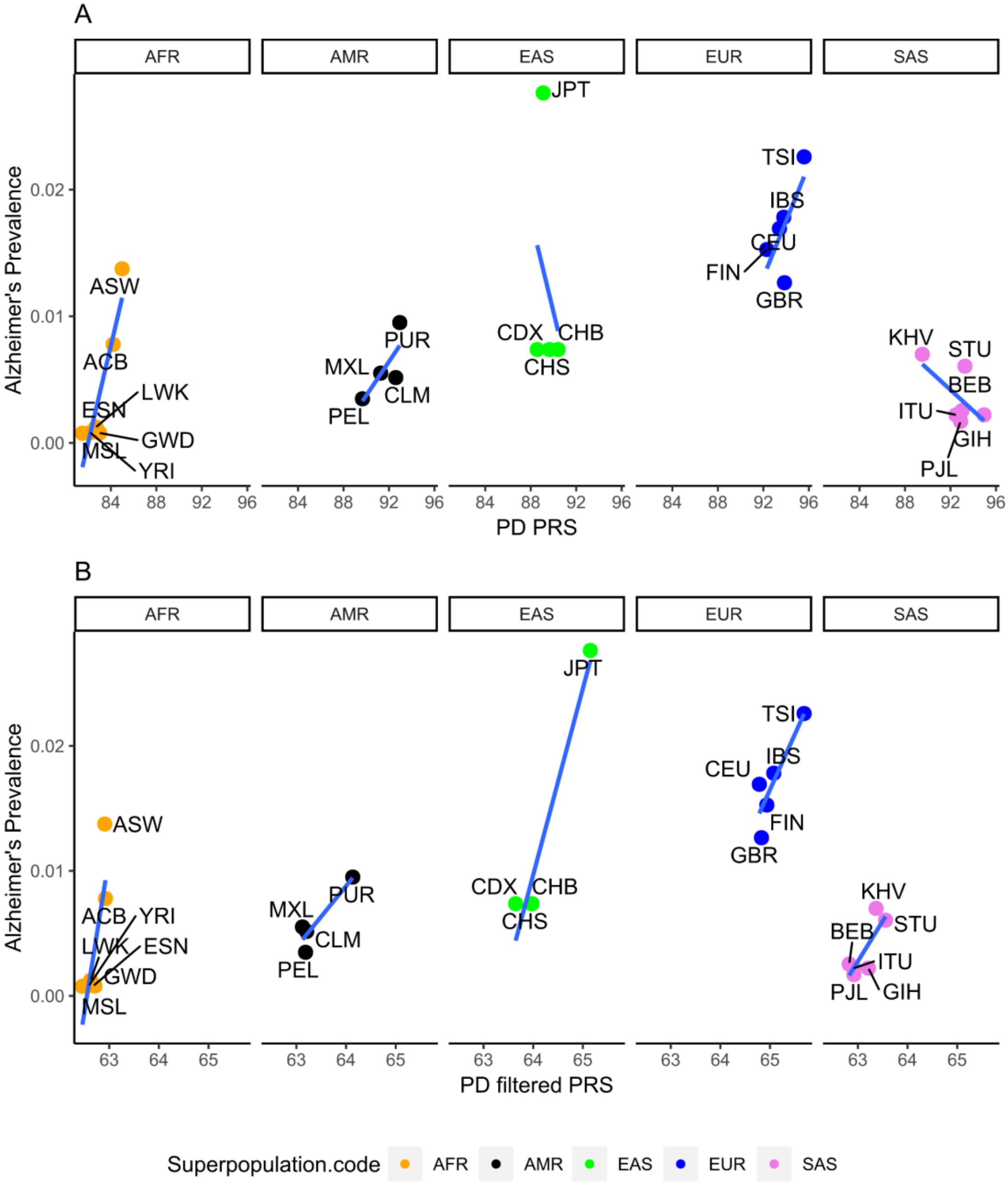


**Figure S4.** PD PRS versus AD prevalence separated by super population. The data points are colored according to the super populations: AFR (orange), AMR (black), EAS (green), EUR (blue) and SAS (purple). A) Full model by super population. B) Super populations after maximization.
